# The impact of long-term conditions and comorbidity patterns on COVID-19 infection and hospitalisation: a cohort study

**DOI:** 10.1101/2023.04.25.23289035

**Authors:** Yun-Ting Huang, Andrew Steptoe, Riyaz S. Patel, Esme Fuller Thomson, Dorina Cadar

## Abstract

**Introduction:** Older adults are usually more vulnerable to COVID-19 infections; however, little is known about which comorbidity patterns are related to a higher probability of COVID-19 infection. This study investigated the role of long-term conditions or comorbidity patterns on COVID-19 infection and related hospitalisations.

**Methods:** This study included 4,428 individuals from Waves 8 (2016−2017) and 9 (2018−2019) of the English Longitudinal Study of Ageing (ELSA), who also participated in the ELSA COVID-19 Substudy in 2020. Comorbidity patterns of chronic conditions were identified using an agglomerative hierarchical clustering method. The relationships between comorbidity patterns or long-term conditions and COVID-19 related outcomes were examined using multivariable logistic regression.

**Results:** Among a representative sample of community-dwelling older adults in England, those with cardiovascular disease (CVD) and complex comorbidities had an almost double risk of COVID-19 infection (OR=1.87, 95% CI=1.42−2.46) but not of COVID-19 related hospitalisation. A similar pattern was observed for the heterogeneous comorbidities cluster (OR=1.56, 95% CI=1.24−1.96). The individual investigations of long-term conditions with COVID-19 infection highlighted primary associations with CVD (OR=1.46, 95% CI=1.23−1.74), lung diseases (OR=1.40, 95% CI=1.17−1.69), psychiatric conditions (OR=1.40, 95% CI=1.16−1.68), retinopathy/eye diseases (OR=1.39, 95% CI=1.18−1.64), and arthritis (OR=1.27, 95% CI=1.09−1.48). In contrast, metabolic disorders and diagnosed diabetes were not associated with any COVID-19 outcomes.

**Discussion/Conclusion:** This study provides novel insights into the comorbidity patterns that are more vulnerable to COVID-19 infections and highlights the importance of CVD and complex comorbidities.

These findings facilitate crucial new evidence for appropriate screening measures and tailored interventions for older adults in the ongoing global outbreak.

## Introduction

COVID-19, also known as SARS-CoV-2, emerged in December 2019 and quickly evolved into a global pandemic, leading to an unprecedented and substantial impact on individuals and nations around the world. Growing evidence suggests that older adults are particularly vulnerable to COVID-19 because they more frequently develop severe forms and complications than younger individuals [1–3].

Current evidence has documented older age, male gender, and pre-existing comorbidities as independent factors associated with poor COVID-19 outcomes: greater severity, higher mortality, and longer hospital stay [4–11]. This has therefore fostered the development of a prediction model for severe COVID-19 [12], in which the importance of comorbidities is emphasised in addition to age and gender. More importantly, atypical COVID-19 symptoms were commonly reported in hospitalised frail older patients, which might have led to difficulty diagnosing this specific age group population [13]. Several mechanisms such as molecular biology, biological ageing, subclinical systemic inflammation, acquired immune response, and ACE2 downregulation (i.e., the SARS-CoV-2 receptor) have been investigated to understand the disproportionate mortality suffered by older adults [1–3]. A recent study points out that phenotypic age (e.g., PhenoAge) predicts adverse COVID-19 outcomes better than chronological age, as it interconnects changes in body composition, energetics, homeostatic mechanisms, and brain health that may contribute to clinical diseases [14].

Several long-term conditions were found to be related to adverse COVID-19 outcomes: acute kidney injury [4, 11], cardiovascular or cerebrovascular disease, respiratory disease (including severe asthma and chronic obstructive pulmonary disease) [5, 11], diabetes [6, 11, 15], hypertension [6], liver disease [8, 11], autoimmune conditions, and history of haematological malignancy or recent other cancer [11] with increased mortality; diabetes [10, 15], chronic lung disease and immunosuppressive conditions [10] with high severity. Nevertheless, other studies present mixed findings on these associations [4, 7]. Other factors such as frailty [4], overweight/obesity [6, 7, 10, 11], shortness of breath [5], deprivation, and non-white ethnicity [11] were also reported as significant risk factors for adverse COVID-19 outcomes.

Furthermore, in addition to individual long-term conditions, a growing number of studies have explored multimorbidity patterns − the coexistence of two long-term conditions. However, which complex comorbidity patterns are related to a higher probability of COVID-19 outcomes is still unclear. Multimorbidity, defined as two or more conditions, is prevalent in adult populations. Cardiometabolic multimorbidity, in particular, has been linked to increased risks of COVID-19 infection [16] and a worse prognosis once infected [17].

Multimorbidity was very common among older adults who had severe COVID-19 infection 1. [18] and among those who died of COVID-19 [19]. The former identified the most common patterns as stroke with hypertension, diabetes and hypertension, and chronic kidney disease and hypertension [18]; the latter study reported hypertension with diabetes, cardiovascular disease, or respiratory disease as the most frequent [19].

To date, the literature has emphasised the severe health consequences of COVID-19 infection rather than assessing the initial risk of contracting COVID-19 among older people. There seems to be controversy over the associations between specific long-term conditions and poor COVID-19 outcomes, with most studies based on those who have already been hospitalised with COVID-19. There also has been limited evidence on comorbidity patterns and COVID-19 outcomes for older adults. This study therefore investigated the associations between long-term conditions or comorbidity patterns and COVID-19 infection and/or related hospitalisation using a representative sample of community-dwelling older adults in England.

## Materials and Methods

### Study population

The data were extracted from Waves 8 (2016−2017) and 9 (2018−2019) of the English Longitudinal Study of Ageing (ELSA), a nationally representative study of adults aged 50 and older living in private households in England [20], and the ELSA COVID-19 Substudy carried out in 2020. The ELSA data collection is carried out every two years using face-to-face computer-assisted interviews followed by self-completion questionnaires. The nurse assessments during which biological samples and anthropometric measurements are taken were completed only on half the samples at Waves 8 and 9, respectively [21]. The ELSA COVID-19 survey was conducted via internet assessment or telephone interviews. In Waves 8 and 9, 10035 people participated in the interviews; 6592 participants were visited by a study nurse. Participants who did not have information on COVID-19 infection, hospitalisation (N=1871), or long-term conditions (N=2) (Table S1) were excluded, resulting in 4719 participants for the cluster analysis presented here. After excluding those who did not have complete information on all selected covariables, 4428 individuals were included as an analytical sample.

### Long-term conditions

This study included all age-related long-term conditions identified in ELSA. Most of them were self-reported by participants but verified by medication use wherever possible. Verified diagnoses of diabetes, lung diseases (including asthma), psychiatric conditions, osteoporosis (including Paget’s disease and heterotopic ossification), Parkinson’s disease, and dementia were employed. In contrast, diagnoses of hyperlipidemia, hypertension, cardiovascular diseases (CVDs), arthritis, cancer, and retinopathy/eye diseases (including diabetic retinopathy, macular degeneration, glaucoma, and cataract) were based on self-reported. A few conditions were defined by recognisably specific treatments, including hyperuricemia (including gout), inflammatory bowel disease and epilepsy. These conditions are primarily supported by the condition list of at-risk groups for COVID-19 on the government website [22]. The association between each condition and infection and/or hospitalisation outcomes was investigated using bivariate analysis. Each significant association was accordingly included in the model as an individual covariate. Diabetes, CVD, lung disease, psychiatric conditions, arthritis, and retinopathy/eye diseases were adjusted separately, while the remaining conditions were included as an illness count in the analyses.

### COVID-19 infection or hospitalisation

Information about COVID-19 infection and related hospitalisation was collected in Waves 1 (June−July 2020) and 2 (November−December 2020) of the COVID-19 Substudy. Participants in the COVID-19 Substudy were recruited from the existing ELSA sample [23]. The ELSA COVID-19 survey adopted a sequential mixed-mode strategy, including computer-assisted web interviewing and computer-assisted telephone interviewing. Participants who reported positive test results for COVID-19 or any of the following COVID-19 symptoms (Wave 1 only) were defined as having COVID-19 infection. The symptoms were high temperature, a new continuous cough, shortness of breath or trouble breathing, fatigue, loss of sense of smell or taste, diarrhoea, abdominal pain, or loss of appetite, published by World Health Organisation (WHO) [24] and National Health Service (NHS) [25]. Hospitalisation due to COVID-19 was defined as having to stay in hospital due to a COVID-19 infection.

### Potential confounders

This study included factors that had been reported in the literature or shown to be significantly related to the outcome in the bivariate analysis. The socio-demographic characteristics included a continuous variable of age (measured in years), a binary variable of sex (male and female), and a categorical variable of total wealth (divided into quintiles). Health factors included self-rated weight (appropriate versus over/underweight, self-reported by participants) and alcohol consumption (i.e., whether a current drinker or not). Features that may influence the rate of COVID-19 infection involved self-isolation and close contact with people who tested positive.

### Statistical analysis

The association between long-term conditions and COVID-19 infection or its hospitalisation was assessed by multivariable logistic regression, and odds ratios (ORs) and 95% confidence intervals (CIs) were reported. Progressive adjustments from the age- and sex-adjusted to the fully-adjusted model were presented. Multivariable logistic regression was also used to assess associations between comorbidity patterns and COVID-19 infection and/or COVID-19 related hospitalisation.

### Cluster analysis

An agglomerative hierarchical clustering approach, with Ward’s linkage and the simple matching coefficient, was employed to group participants by taking account of similarity among the 15 long-term conditions [26–28]. Agglomerative hierarchical methods begin with each observation in its own group; then, the two closest (most similar) groups are combined (all the rest of the groups remain single), which is done repeatedly until the desired number of clusters is reached. Ward’s linkage method is also known as the minimum sum of squares, in which the lowest sum of squared distances is chosen to be combined [26]. The simple matching coefficient is the most common method when binary data is used in cluster analysis [27]. Considering the small number of ELSA participants who were hospitalised due to COVID-19 (N=27; Table S2), a maximum of 10 clusters were considered in order to maintain sufficient statistical power in each cluster. The dendrogram of cluster analysis is shown in Figure S1. The results indicated that four clusters fitted best the data, with lower Akaike information criterion (AIC) and Bayesian information criterion (BIC) (Table S3). The clusters were labelled based on the most prevalent condition, incorporating each cluster’s comorbidity pattern. The statistical analyses were conducted using Stata (version 15.1; StataCorp LP, College Station, TX, USA).

### Sensitivity and supplementary analysis

Two sensitivity analyses and a supplementary analysis were performed to ensure the robustness of the main findings. The first sensitivity analysis further stratified diabetes into several categories: no diabetes, diagnosed diabetes with and without metformin use, and undiagnosed diabetes, defined as not having self-reported diabetes and any diabetic medications but having a glycated haemoglobin measurement ≥ 48 mmol/mol (equivalent to 6.5%). The second sensitivity analysis employed a stricter definition of COVID-19 infection, including only three main symptoms (i.e. high temperature, a new continuous cough, and loss of sense of smell or taste) according to the UK NHS [25] and shortness of breath or trouble breathing, which is a typical symptom of pneumonia. Lastly, a supplementary analysis was carried out to examine the baseline characteristics between complete cases and samples with missing values.

## Results

The baseline characteristics of the study sample are summarised in Table 1. The mean age of the sample was 69.2 years, and 56.6% were women. Approximately half were classified as having higher wealth (e.g., in the upper two quintiles of wealth). Arthritis was the most prevalent long-term condition (43.6%), followed by retinopathy/eye diseases (40.8%) and CVDs (22.4%). Over half were current drinkers according to self-report (63.4%) or over/underweight (58.0%). During the pandemic, 32.5% reported having self-isolation, and 13.8% had close contact with someone who tested positive. A total of 21.8% had COVID-19 infection, but only 0.6% were admitted to hospitals.

**Table 1.** Baseline characteristics among 4428 individuals, ELSA 2016/2018.

|  | % (N) |
| --- | --- |
| Age (years) mean (SD) | 69.2 (7.9) |
| Women | 56.6 (2506) |
| Total wealth |  |
| 1 (lowest) | 12.5 (553) |
| 2 | 15.9 (705) |
| 3 | 22.5 (998) |
| 4 | 24.5 (1084) |
| 5 (highest) | 24.6 (1088) |
| Diabetes | 13.1 (580) |
| CVD | 22.4 (993) |
| Lung disease | 17.1 (758) |
| Psychiatric conditions | 18.3 (810) |
| Arthritis | 43.6 (1929) |
| Retinopathy/eye diseases | 40.8 (1807) |
| Number of conditions median (IQR) | 1 (2) |
| Current drinker | 63.4 (2807) |
| Self-rated over- or underweight | 58.0 (2567) |
| Self-isolation* | 32.5 (1441) |
| Close contact with people who tested positive* | 13.8 (609) |
| COVID-19 infection*† | 21.8 (965) |
| Hospitalisation due to COVID-19 infection* | 0.6 (27) |

\* Data was collected in ELSA COVID-19 Substudy
† Including people who self-reported COVID-19-like symptoms at wave 1 of ELSA COVID-19 Substudy

### Long-term conditions and COVID-19 infection/hospitalisation

The results of associations between long-term conditions and COVID-19 infection or its hospitalisation are shown in Table 2. CVDs (OR=1.46, 95% CI=1.23−1.74), lung diseases (OR=1.40, 95% CI=1.17−1.69), psychiatric conditions (OR=1.40, 95% CI=1.16−1.68), retinopathy/eye diseases (OR=1.39, 95% CI=1.18−1.64), and arthritis (OR=1.27, 95% CI=1.09−1.48) were strongly associated with COVID-19 infection, with slight attenuation of ORs from the age- and gender-adjusted model to the fully adjusted model. Diabetes was not related to COVID-19 infection, but those with diabetes had a lower risk of hospitalisation due to COVID-19 infection (OR=0.10, 95% CI=0.01−0.80). Those with retinopathy/eye diseases had a 3.38 times higher risk of hospitalisation, while other conditions did not show any association.

**Table 2.** Associations* between long-term conditions and COVID-19 infection or its hospitalisation (N=4428), England 2016/2018−2020.

|  | Model 1: age and gender |  | Model 2: model 1 + wealth |  | Model 3: model 2 +<br>lifestyle factors<sup>†</sup> |  | Model 4: model 3 +<br>COVID-19-related factors<sup>#</sup> |  |
| --- | --- | --- | --- | --- | --- | --- | --- | --- |
| **COVID-19 infection** | OR (95% CIs) | P | OR (95% CIs) | P | OR (95% CIs) | P | OR (95% CIs) | P |
| Diabetes | 1.03 (0.84, 1.28) | 0.754 | 1.03 (0.83, 1.28) | 0.794 | 1.00 (0.80, 1.24) | 0.969 | 0.99 (0.80, 1.24) | 0.959 |
| CVD | 1.50 (1.26, 1.78) | **<0.001** | 1.49 (1.26, 1.77) | **<0.001** | 1.49 (1.25, 1.77) | **<0.001** | 1.46 (1.23, 1.74) | **<0.001** |
| Lung disease | 1.48 (1.24, 1.78) | **<0.001** | 1.48 (1.23, 1.77) | **<0.001** | 1.47 (1.23, 1.76) | **<0.001** | 1.40 (1.17, 1.69) | **<0.001** |
| Psychiatric conditions | 1.43 (1.20, 1.72) | **<0.001** | 1.42 (1.19, 1.71) | **<0.001** | 1.41 (1.17, 1.69) | **<0.001** | 1.40 (1.16, 1.68) | **<0.001** |
| Arthritis | 1.30 (1.12, 1.52) | **0.001** | 1.30 (1.11, 1.51) | **0.001** | 1.28 (1.10, 1.49) | **0.002** | 1.27 (1.09, 1.48) | **0.003** |
| Retinopathy/eye diseases | 1.40 (1.19, 1.65) | **<0.001** | 1.40 (1.19, 1.65) | **<0.001** | 1.40 (1.19, 1.64) | **<0.001** | 1.39 (1.18, 1.64) | **<0.001** |
| Number of conditions<sup>§</sup> | 1.08 (0.99, 1.17) | 0.073 | 1.07 (0.99, 1.16) | 0.084 | 1.07 (0.98, 1.16) | 0.116 | 1.06 (0.98, 1.15) | 0.162 |
| **Hospitalisation due to**<br>**COVID-19 infection** | OR (95% CIs) | P | OR (95% CIs) | P | OR (95% CIs) | P | OR (95% CIs) | P |
| Diabetes | 0.14 (0.02, 1.08) | 0.060 | 0.12 (0.02, 0.91) | **0.040** | 0.11 (0.01, 0.83) | **0.032** | 0.10 (0.01, 0.80) | **0.030** |
| CVD | 1.64 (0.72, 3.69) | 0.236 | 1.52 (0.67, 3.42) | 0.314 | 1.51 (0.67, 3.41) | 0.324 | 1.44 (0.64, 3.26) | 0.378 |
| Lung disease | 1.91 (0.84, 4.36) | 0.125 | 1.83 (0.80, 4.19) | 0.150 | 1.84 (0.80, 4.22) | 0.149 | 1.57 (0.67, 3.65) | 0.295 |
| Psychiatric conditions | 1.07 (0.41, 2.75) | 0.891 | 0.89 (0.34, 2.32) | 0.805 | 0.85 (0.32, 2.23) | 0.734 | 0.84 (0.32, 2.21) | 0.718 |
| Arthritis | 1.30 (0.58, 2.94) | 0.522 | 1.16 (0.51, 2.63) | 0.723 | 1.13 (0.49, 2.59) | 0.775 | 1.11 (0.48, 2.54) | 0.813 |
| Retinopathy/eye diseases | 3.38 (1.32, 8.69) | **0.011** | 3.41 (1.32, 8.81) | **0.011** | 3.50 (1.35, 9.09) | **0.010** | 3.38 (1.30, 8.82) | **0.013** |
| Number of conditions<sup>§</sup> | 1.43 (0.98, 2.09) | 0.065 | 1.34 (0.92, 1.96) | 0.128 | 1.34 (0.92, 1.96) | 0.128 | 1.28 (0.87, 1.87) | 0.208 |

\* Adjusted for age, gender, total wealth, alcohol consumption (a current drinker or not), self-rated weight, self-isolation, and close contact with people who tested positive
† Including alcohol consumption and self-rated weight

### Comorbidity patterns and COVID-19 infection/hospitalisation

Four clusters (comorbidity patterns) were identified based on the 15 long-term conditions. The distribution of long-term conditions across the four clusters is displayed in Figure 1.

- Cluster 1 comprised 1389 individuals, of whom over half were diagnosed with hyperlipidemia (69.6%) and hypertension (55.4%). This cluster also had the highest prevalence of diabetes (32.5%), and it, therefore, was labelled ‘Metabolic disorders’.
- Cluster 2 consisted of 1666 participants who were frequently diagnosed with retinopathy/eye diseases (57.3%) and arthritis (47.4%), which were also prevalent in other clusters. This cluster had the highest prevalence of lung diseases (33.2%), psychiatric conditions (29.4%), osteoporosis (21.6%), and cancer (18.8%) among all clusters. Cluster 2 was labelled ‘heterogeneous comorbidities’ due to a lack of specific comorbidity patterns.
- Cluster 3 comprised 596 individuals who had CVDs (100.0%), with 59.7% hypertension, 56.5% arthritis, 54.5% retinopathy/eye diseases, and 48.3% hyperlipidemia. As a result, this cluster was labelled ‘CVD with complex comorbidities’.
- Cluster 4 consisted of 777 participants, of whom 27.0% had arthritis, with a very low prevalence of other conditions (< 3%). It was therefore labelled ‘Healthiest’.

**Figure 1.**
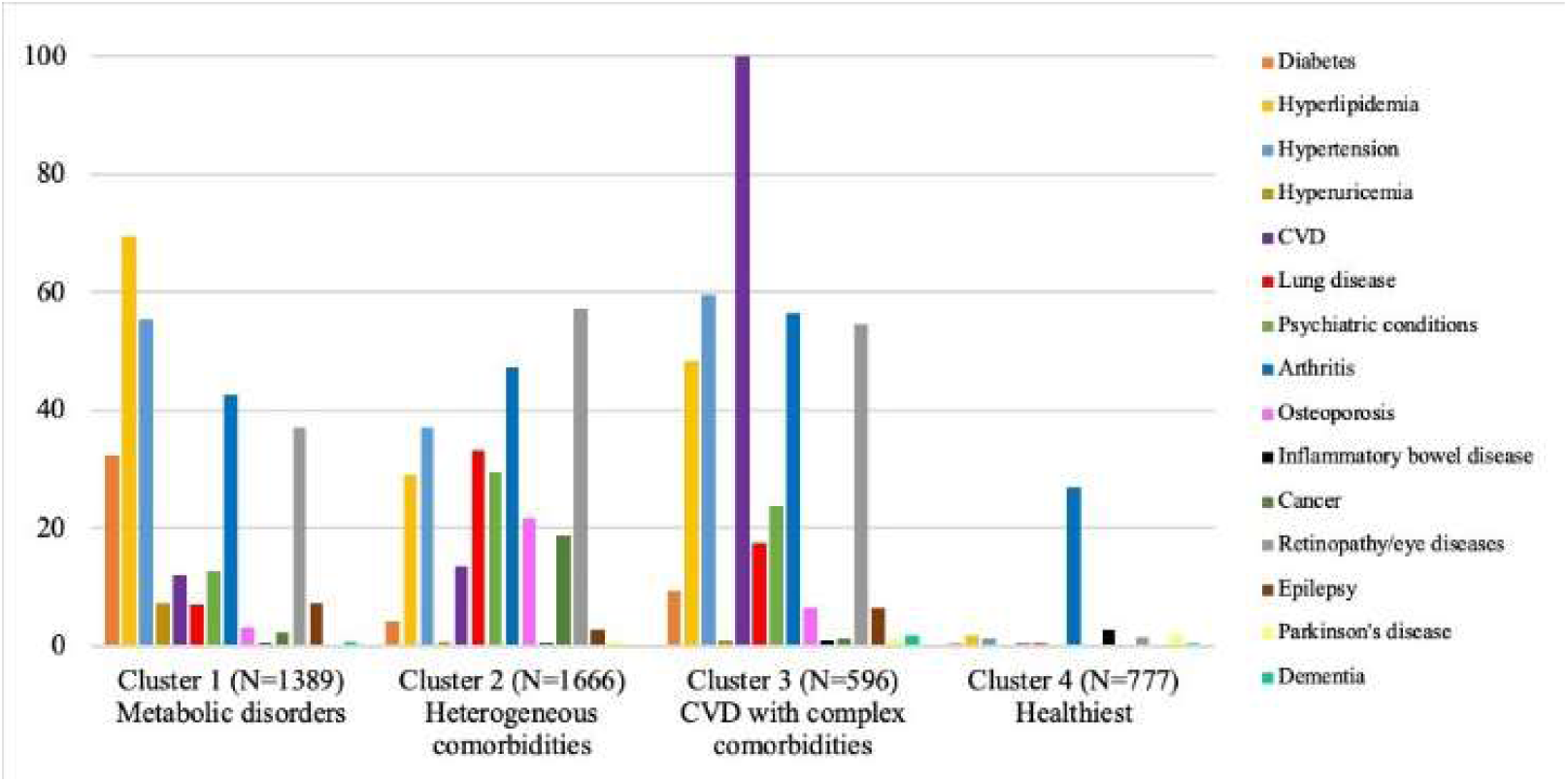
Prevalence of long-term conditions across clusters, ELSA 2016/2018.

Table 3 shows the results of the association between pre-pandemic comorbidity patterns (at baseline in 2016/2018) and COVID-19 infection and/or related hospitalisation (in 2020). The CVD with complex comorbidities cluster showed an 87% increase in the risk of COVID-19 infection (OR=1.87, 95% CI=1.42−2.46), but not of hospitalisation compared with the healthiest cluster. A similar pattern was observed for the heterogeneous comorbidities cluster, with a more minor increase of 56% (OR=1.56, 95% CI=1.24−1.96), whereas the metabolic disorders cluster did not reveal any association with either COVID-19 outcomes. The CVD with complex comorbidities cluster and heterogeneous comorbidities cluster also had a higher prevalence of lung diseases (17.6% and 33.2%) and psychiatric conditions (23.8% and 29.4%) than the metabolic disorders cluster and healthiest cluster.

Apart from comorbidity patterns, respondents who were over/underweight (OR=1.21, 95% CI=1.04−1.41) and those who had close contact with people who tested positive for COVID-19 (OR=1.57, 95% CI=1.29−1.91) had a higher risk of COVID-19 infection. In contrast, those who were older were less likely to become infected (OR=0.98, 95% CI=0.97−0.99). Only two factors were significantly associated with hospitalisation due to COVID-19 infection: with every increase in the quintile of wealth, the risk of hospitalisation was lower by 36% (OR=0.64, 95% CI=0.47−0.87), and those who had self-isolation during the pandemic showed double the risk of subsequent COVID-19 hospitalisation (OR=2.62, 95% CI=1.13−6.12).

**Table 3.** Associations between comorbidity patterns and COVID-19 infection or its hospitalisation (N=4428), England 2016/2018−2020.

|  | COVID-19 infection |  | Hospitalisation due to COVID-19 infection |  |
| --- | --- | --- | --- | --- |
|  | OR (95% CIs) | P | OR (95% CIs) | P |
| Comorbidity pattern * |  |  |  |  |
| Metabolic disorders (cluster 1) | 1.13 (0.89, 1.43) | 0.326 | 1.51 (0.17, 13.81) | 0.715 |
| Heterogeneous comorbidities (cluster 2) | 1.56 (1.24, 1.96) | **<0.001** | 4.32 (0.55, 33.71) | 0.163 |
| CVD with complex comorbidities (cluster 3) | 1.87 (1.42, 2.46) | **<0.001** | 4.48 (0.53, 38.14) | 0.170 |
| Age (years) | 0.98 (0.97, 0.99) | **0.001** | 1.02 (0.97, 1.08) | 0.343 |
| Women | 1.04 (0.90, 1.21) | 0.581 | 0.67 (0.30, 1.48) | 0.326 |
| Total wealth (quintiles) | 0.97 (0.91, 1.02) | 0.228 | 0.64 (0.47, 0.87) | **0.005** |
| Current drinker | 0.86 (0.73, 1.00) | 0.053 | 0.68 (0.30, 1.52) | 0.350 |
| Self-rated over- or underweight | 1.21 (1.04, 1.41) | **0.013** | 0.87 (0.39, 1.91) | 0.721 |
| Self-isolation | 1.39 (1.19, 1.63) | **<0.001** | 2.62 (1.13, 6.12) | **0.025** |
| Close contact with people who tested positive | 1.57 (1.29, 1.91) | **<0.001** | 1.84 (0.72, 4.68) | 0.202 |

\* Cluster 4 Healthiest as the reference group

### Sensitivity and supplementary analyses

As shown in Table S4, individuals with undiagnosed diabetes before the pandemic had almost five times higher risk of COVID-19 related hospitalisation (OR=4.80, 95% CI=1.01−22.73) than those without diabetes and elevated glycated haemoglobin levels at baseline. The use of metformin was found not to be related to COVID-19 infection or its hospitalisation. After metformin intake was accounted for, diabetes was no longer associated with COVID-19 hospital admissions. No diabetes patient who did not take metformin was hospitalised due to COVID-19 infection, leading to a failure estimation in the model. The second sensitivity analysis with a stricter definition of COVID-19 infection demonstrated similar results (Table S5) with the primary results, confirming the robustness of this study. The CVD with complex comorbidities cluster (OR=1.86, 95% CI=1.32−2.61) and heterogeneous comorbidities cluster (OR=1.44, 95% CI=1.07−1.93) were consistently associated with a high risk of COVID-19 infection. The positive relationships between CVD (OR=1.47, 95% CI=1.19−1.82), lung disease (OR=1.72, 95% CI=1.38−2.13), or retinopathy/eye diseases (OR=1.36, 95% CI=1.10−1.67) and COVID-19 infection remained significant, whilst the association with psychiatric conditions and arthritis disappeared, probably attributable to lower power. Lastly, the distribution of baseline characteristics was similar between complete cases and samples with missingness (Table S6).

## Discussion

Among community-dwelling adults aged 50 and older in England, those with CVD and complex comorbidities and those with heterogeneous comorbidities had an 87% and 56% increase in the risk of COVID-19 infection, respectively, but not in COVID-19 related hospitalisation. The metabolic disorders cluster had no association with either COVID-19 associated outcomes. Several pre-existing long-term conditions before the pandemic, including CVDs, lung diseases, psychiatric disorders, retinopathy/eye diseases, and arthritis, were found to be related to the risk of subsequent COVID-19 infection. Older adults with retinopathy/eye diseases were also at a high risk of COVID-19 related hospital admissions. After accounting for metformin use and undiagnosed diabetes, those with undiagnosed diabetes had a higher risk of COVID-19 related hospitalisation, while diagnosed diabetes was no longer associated with COVID-19 hospital admissions. The robustness of this study was primarily supported by sensitivity and supplementary analyses. The similar distribution of baseline characteristics between complete cases and samples with missing values indicated that missingness is less likely to lead to selection bias.

The disparity in the prevalence of three long-term conditions may account for the differential risks of COVID-19 infection among different comorbidity patterns. Lung diseases and psychiatric disorders were independently associated with COVID-19 infection and more prevalent in the clusters of CVD with complex comorbidities (17.6% and 23.8%) and heterogeneous comorbidities (33.2% and 29.4%) than in the metabolic disorders cluster (7.0% and 12.7%). In contrast, each former cluster was less likely to coexist with diabetes (9.4% and 4.1%), found not to be related to COVID-19 infection, than the metabolic disorders cluster (32.5%). The metabolic disorders cluster also showed a low prevalence of CVD (12.0%), apart from lung diseases and psychiatric conditions, and this might account for the lack of association with COVID-19 infection. To our knowledge, this study is the first to investigate the associations between complex comorbidity patterns (beyond multimorbidity patterns) and COVID-19 infection and subsequent hospital admissions among older adults; thus, direct comparisons with previous studies are difficult to make. Cardiometabolic multimorbidity reportedly had a high risk of COVID-19 infection and worse outcomes in adults [16, 17] and older adults [18, 19], partially supporting our findings on CVD with complex comorbidities.

CVDs, lung diseases, and psychiatric conditions are regarded as having a high risk of severe illness from COVID-19 according to NHS [22] and the Centers for Disease Control and Prevention (CDC) websites [29] and previous studies [5, 10, 11]. Older adults with such long-term conditions may develop symptoms when they contract COVID-19, rather than being asymptomatic. Therefore, they may be more likely to get tested than their peers without these conditions. This study included participants with relevant symptoms at Wave 1 as COVID-19 cases, apart from those who reported positive COVID-19 test results, since the testing capacity in the UK had not been sufficient at the time data was collected. Future research would benefit from universal screening for COVID-19 rather than only testing symptomatic individuals. The pathophysiological mechanism of pre-existing CVDs and COVID-19 infection is still unclear, although some potential direct SARS-CoV-2 and indirect immune response mechanisms impacting the cardiovascular system have been explored and discussed [30]. There is strong evidence to suggest that older adults with lung diseases are likely to be symptomatic or develop severe symptoms because COVID-19 targets the respiratory system, including the lungs. Individuals with psychiatric conditions may have a higher chance of living in crowded residences or working in unsafe environments where infections can spread quickly, due to increased levels of socioeconomic deprivation. This group of people would inevitably become highly vulnerable to COVID-19 infection [31].

Both arthritis and retinopathy/eye diseases were prevalent across different comorbidity patterns, so it might be challenging to conclude their impact on the risk of COVID-19 infection. Arthritis is a heterogeneous disease that may refer to rheumatoid arthritis, psoriatic arthritis, osteoarthritis etc. Different types of arthritis have diverse underlying disease mechanisms and therefore need different treatments. Given the lack of information about the arthritis types, it would be daring to interpret the potential effects of different treatments, such as non-steroidal anti-inflammatory drugs (NSAIDs) or disease-modifying antirheumatic drugs (DMARDs), on the risk of COVID-19 infection. The heterogeneous comorbidities, however, showed the highest prevalence of osteoporosis in the combination of a high proportion of arthritis, implying that a certain number of people in this cluster might have autoimmune diseases (e.g. rheumatoid arthritis and psoriatic arthritis) because osteoporosis is linked to the malfunctioning immune system [32]. Autoimmune diseases often require DMARDs to suppress overactive immune and/or inflammatory systems, so this immunosuppression may increase the risk of being infected with COVID-19. People with autoimmune diseases have been found to have an increased risk of getting infected with COVID-19, but not more severe, which is closely related to underlying inflammation and pharmacological treatments of autoimmune diseases [33].

The retinopathy/eye diseases in this study included four age-related conditions: diabetic retinopathy, macular degeneration, glaucoma, and cataract, and they are closely or partially related to inflammation [34–37] that might influence the risk of COVID-19 infection. Diabetic retinopathy was found to be independently associated with severe COVID-19 infection requiring intensive care treatment, although the mechanism is uncertain [38]. Older adults with arthritis or retinopathy/eye diseases are more likely to develop limitations in activities of daily living and become more dependent. Higher dependency would increase their exposure to personal support workers and other health care professionals, which may increase the likelihood of exposure to COVID-19.

Older adults diagnosed with diabetes in the ELSA are likely to manage this condition well because there are many NHS support groups available throughout the country, and ELSA, as a longitudinal study, provides participants with feedback on their blood biomarkers. This may account for the lack of association between diagnosed diabetes and COVID-19 outcomes in this study, in line with some previous research [4, 7]. In contrast, several other studies concluded that diabetes was a risk factor for adverse outcomes of COVID-19 infection [6, 10, 11, 15]. The discrepancy between our findings and most of the literature might also be attributed to a high proportion of wealthy participants in our study when deprivation was thought to contribute to poor COVID-19 outcomes. In addition, this study found that participants with undiagnosed diabetes had an increased risk of COVID-19 hospitalisation, supported by a Mexican study [39]. The elevated plasma glucose level was found to play an important role in mediating the effect of acute inflammation on adverse COVID-19 outcomes [39] and might contribute to the positive association between undiagnosed diabetes and COVID-19 hospitalisation. Future investigations in larger samples of COVID-19 hospitalisation are needed to confirm our findings on diagnosed and undiagnosed diabetes. However, our study highlights the importance of diabetes screening and management among older adults.

Among other factors investigated, older age and male gender have been identified as risk factors for poor COVID-19 outcomes in many earlier studies [4–11]; however, older people were less likely to be infected or symptomatic in this study, with no gender difference observed. ELSA participants aged 65 and older might not need to work, so they would have a lower chance of contracting COVID-19 than those who went to work. Wealthier people had a lower risk of COVID-19 hospital admissions, in line with previous findings of deprivation and COVID-19 deaths [11], despite no significant difference in the risk of being infected. This study’s finding that self-rated over/underweight was a risk factor associated with COVID-19 infection is in keeping with previous research linking obesity to adverse COVID-19 outcomes [6, 7, 10, 11]. Older adults who had experienced self-isolation during the pandemic were more likely to be infected or hospitalised due to COVID-19, although this could also represent reverse causation. Furthermore, these could have been individuals who were clinically extremely vulnerable as suggested by the UK government criteria and then advised to shield themselves during earlier stages of the pandemic, those who had any symptoms of COVID-19 or felt unwell, those who had tested positive for COVID-19, and those who had been in close contact with someone with COVID-19, which would reasonably increase the risk. After we accounted for self-isolation, we found that close contact with someone who tested positive was related to the risk of COVID-19 but not of COVID-19 hospitalisation.

### Strengths and limitations

This novel study has several important strengths. First, although health conditions were originally identified through self-report by participants, most of these conditions were verified through the medication profiles collected by nurses. Nurses recorded medication data according to the containers of the prescribed medicines the participant took during home visits. This triangulation method using medication verification and a reliable collection process helped reduce reporting bias. Second, the study employed a nationally representative sample, for whom comprehensive pre-pandemic characteristics were available ranging from socio-demographic characteristics to health status. Third, we adjusted for a wider range of potential confounders, including factors that may contribute to COVID-19 infection collected in the ELSA COVID-19 Substudy, than those addressed in most previous research. Lastly, the adoption of cluster analysis – an advanced statistical technique – allows researchers to take coexisting long-term conditions into account in a rich and nuanced way. The clustering approach groups older people into clusters based on their long-term conditions, regardless of their individual risk of COVID-19, because the coexistence of multiple conditions should not be influenced by COVID-19 infection. This methodological strategy is data-driven and different from traditional analyses, which often treat each condition as a covariate and/or which rely on researchers to define multimorbidity patterns. Coexisting long-term conditions are common among older adults and may interact with each other, further influencing the risk of COVID-19 outcomes.

Some limitations of this study should also be acknowledged. Most of the information was self-reported, so recall bias could not be avoided. The inclusion of multiple COVID-19 symptoms based on WHO criteria may have led to misclassification bias since these symptoms are not unique to COVID-19 infection and may have been attributed to other respiratory tract infections (e.g., flu). Participants in our study seemed to be less deprived, which may have hampered the representativeness of this study. Lastly, the lack of significant associations between the comorbidity patterns and COVID-19 hospitalisation might be due to low statistical power, attributable to the small number of hospital admissions recorded in this study.

### Implications for clinical practice

Research understanding of COVID-19 is evolving with a particular need for further understanding older adults’ needs. The guidance published by the CDC and NHS identifies those with several medical conditions (e.g., cancer, chronic lung diseases, and heart conditions) as having a higher risk of severe illness from COVID-19 [29, 40]. However, it seems less clear what types of comorbidity patterns would place older adults at higher risk of being infected or hospitalised for COVID-19. The results of this study provide new evidence on the link between comorbidity patterns and the risk of COVID-19 infection and subsequent hospitalisation amongst older adults and highlight the importance of CVD with complex comorbidities that should be paid more attention to. This study is expected to reinforce the need for tailored public health policies that evolve with the COVID-19 pandemic to improve prevention strategies such as self-isolation policy and regular booster vaccinations, and even treatment strategies (e.g., early administration of antiviral medications) for vulnerable ageing populations.

## Conclusion

Among community-dwelling older adults in England, those who had CVD and complex comorbidities had an almost double risk of COVID-19 infection. This was not the case for those with metabolic disorders or diagnosed diabetes, but those with undiagnosed diabetes might present a higher risk of COVID-19 hospitalisation. This study provides novel evidence on the increased risk of COVID-19 infection among older adults with complex comorbidity patterns and reiterates the need for personalised screening and public health interventions such as regular booster vaccinations for those with CVD and complex comorbidities.

## Acknowledgements

The authors would like to thank NatCen (National Centre for Social Research), the interviewers, the nurses, and the researchers involved in the data collection process. Also, the authors are grateful to all ELSA participants for their commitment to the study and contribution.

## Statement of Ethics

ELSA has received ethical approval from the South Central – Berkshire Research Ethics Committee (21/SC/0030, 22^nd^ March 2021). Written informed consent for this particular study was not required.

## Author Contributions

Yun-Ting Huang conceived and designed the study and carried out the analysis. Dorina Cadar advised on the study design and policy implications. Yun-Ting Huang and Dorina Cadar verified the underlying data. All authors contributed to the interpretation of the findings. Dorina Cadar, Esme Fuller Thomson, Andrew Steptoe and Riyaz S. Patel were responsible for securing the funding. Yun-Ting Huang drafted the initial manuscript, and all authors contributed to the revisions of this manuscript. All authors read and approved the final manuscript.

## Data Availability Statement

The datasets generated and/or analysed during the current study are not publicly available due to ethical reasons. Further enquiries can be directed to the corresponding author. Researchers can download ELSA data from all waves from the UK Data Service.

