## Supplementary materials for "The impact of long-term conditions and comorbidity patterns on COVID-19 infection and hospitalisation: a cohort study"

**Table S1. Prevalence of long-term conditions among 4428 individuals, ELSA 2016/2018**

| **Long-term conditions** | **% (N)** |
|---|---|
| Arthritis | 43.6 (1929) |
| Retinopathy/eye diseases (including diabetic retinopathy, macular degeneration, glaucoma, and cataract) | 40.8 (1807) |
| Hyperlipidemia | 39.5 (1750) |
| Hypertension | 39.5 (1751) |
| CVD | 22.4 (993) |
| Psychiatric conditions† | 18.3 (810) |
| Lung disease† | 17.1 (758) |
| Diabetes† | 13.1 (580) |
| Osteoporosis (including Paget's disease and heterotopic ossification)† | 10.0 (443) |
| Cancer | 8.0 (354) |
| Epilepsy# | 4.3 (188) |
| Hyperuricemia (including gout) # | 2.8 (122) |
| Inflammatory bowel disease# | 1.0 (42) |
| Parkinson's disease† | 0.9 (39) |
| Dementia† | 0.8 (33) |

† Verified diagnosis
### Defined by recognisably specific treatments

**Table S2. Rates of COVID-19 infection or its hospitalisation by cluster, ELSA COVID-19 Substudy in 2020**

| | **Cluster 1**<br>**Metabolic disorders**<br><br>(N=1389)<br>% (N) | **Cluster 2**<br>**Heterogeneous comorbidities**<br><br>(N=1666)<br>% (N) | **Cluster 3**<br>**CVD with complex comorbidities**<br><br>(N=596)<br>% (N) | **Cluster 4**<br>**Healthiest**<br><br>(N=777)<br>% (N) |
|---|---|---|---|---|
| COVID-19 infection | 18.7 (260) | 24.4 (406) | 28.2 (168) | 16.9 (131) |
| Hospitalisation due to COVID-19 infection | 0.3 (4) | 0.9 (15) | 1.2 (7) | 0.1 (1) |

**Table S3. Model fit statistics for other cluster options (N=4428)**

| | 2-cluster | 3-cluster | 4-cluster | 5-cluster | 6-cluster |
|---|---|---|---|---|---|
| Sample size in each cluster | 3651, 777 | 3055, 596, 777 | 1389, 1666, 596, 777 | 425, 964, 1666, 596, 777 | 425, 964, 384, 1282, 596, 777 |
| COVID-19 infection | | | | | |
| Log likelihood | -2278.6364 | -2273.8478 | -2267.5284 | -2266.036 | -2264.6177 |
| AIC | 4575.273 | 4567.696 | 4557.057 | 4556.072 | 4555.235 |
| BIC | 4632.834 | 4631.653 | 4627.41 | 4632.82 | 4638.379 |
| Hospitalisation due to COVID-19 infection | | | | | |
| Log likelihood | -149.66496 | -149.35208 | -147.35708 | -147.18351 | -145.6499 |
| AIC | 317.3299 | 318.7042 | 316.7142 | 318.367 | 317.2998 |
| BIC | 374.8912 | 382.6612 | 387.0669 | 395.1155 | 400.4439 |

**Table S4. Sensitivity analyses of the associations* between metformin use/undiagnosed diabetes and COVID-19 infection or its hospitalisation (N=4428), England 2016/2018−2020**

| | **COVID-19 infection** | | **Hospitalisation due to COVID-19 infection#** | |
|---|---|---|---|---|
| | OR (95% CIs) | P | OR (95% CIs) | P |
| Diabetes† | | | | |
| Diagnosed with metformin | 0.89 (0.67, 1.18) | 0.416 | 0.19 (0.02, 1.46) | 0.109 |
| Diagnosed without metformin | 1.13 (0.83, 1.52) | 0.437 | 1.00 | — |
| Undiagnosed | 0.78 (0.37, 1.68) | 0.532 | 4.80 (1.01, 22.73) | **0.048** |
| CVD | 1.46 (1.23, 1.74) | **<0.001** | 1.45 (0.64, 3.30) | 0.371 |
| Lung disease | 1.41 (1.17, 1.69) | **<0.001** | 1.52 (0.65, 3.55) | 0.337 |
| Psychiatric conditions | 1.40 (1.16, 1.68) | **<0.001** | 0.84 (0.32, 2.22) | 0.725 |
| Arthritis | 1.26 (1.08, 1.48) | **0.003** | 1.13 (0.49, 2.58) | 0.781 |
| Retinopathy/eye diseases | 1.39 (1.18, 1.64) | **<0.001** | 3.34 (1.28, 8.69) | **0.013** |
| Number of conditions§ | 1.06 (0.98, 1.15) | 0.154 | 1.25 (0.85, 1.84) | 0.251 |

* Adjusted for age, gender, total wealth, alcohol consumption (a current drinker or not), self-rated weight, self-isolation, and close contact with people who tested positive

† 330 participants with diagnosed diabetes and metformin; 250 participants with diagnosed diabetes but without metformin; 43 participants with undiagnosed diabetes

### Of 27 individuals who were hospitalised, 24 had no diabetes; 1 had diagnosed diabetes and took metformin; 2 had undiagnosed diabetes

§ The other remaining conditions, not including diabetes, CVD, lung disease, psychiatric conditions, arthritis, and retinopathy/eye diseases

**Table S5. Sensitivity analyses of the associations between long-term conditions or comorbidity patterns and COVID-19 infection, with a stricter definition (N=4428), England 2016/2018−2020**

| | **COVID-19 infection** | |
|---|---|---|
| Single long-term conditions† | OR (95% CIs) | P |
| Diabetes | 0.87 (0.66, 1.15) | 0.329 |
| CVD | 1.47 (1.19, 1.82) | **<0.001** |
| Lung disease | 1.72 (1.38, 2.13) | **<0.001** |
| Psychiatric conditions | 1.14, (0.91, 1.44) | 0.251 |
| Arthritis | 1.20 (0.99, 1.46) | 0.069 |
| Retinopathy/eye diseases | 1.36 (1.10, 1.67) | **0.004** |
| Number of conditions§ | 1.09 (0.98, 1.20) | 0.097 |
| Comorbidity pattern* | OR (95% CIs) | P |
| Metabolic disorders (cluster 1) | 1.01 (0.74, 1.38) | 0.949 |
| Heterogeneous comorbidities (cluster 2) | 1.44 (1.07, 1.93) | **0.015** |
| CVD with complex comorbidities (cluster 3) | 1.86 (1.32, 2.61) | **<0.001** |

† Fully adjusted model (Model 4)
* Cluster 4 Healthiest as the reference group

**Table S6. Baseline characteristics between complete cases (N=4428) and samples with missing values, ELSA 2016/2018**

| | Complete cases (N=4428) | Sample with missing values | |
|---|---|---|---|
| | % (N) | % (N) | Sample size |
| Age (years) mean (SD) | 69.2 (7.9) | 69.2 (8.0) | 4690 |
| Women | 56.6 (2506) | 56.3 (2657) | 4720 |
| Total wealth | | | |
| 1 (lowest) | 12.5 (553) | 12.9 (597) | 4637 |
| 2 | 15.9 (705) | 16.0 (742) | |
| 3 | 22.5 (998) | 22.5 (1044) | |
| 4 | 24.5 (1084) | 24.4 (1129) | |
| 5 (highest) | 24.6 (1088) | 24.3 (1125) | |
| Diabetes | 13.1 (580) | 13.2 (621) | 4720 |
| CVD | 22.4 (993) | 22.4 (1057) | 4720 |
| Lung disease | 17.1 (758) | 17.2 (811) | 4720 |
| Psychiatric conditions | 18.3 (810) | 18.1 (854) | 4720 |
| Arthritis | 43.6 (1929) | 43.6 (2060) | 4720 |
| Retinopathy/eye diseases | 40.8 (1807) | 40.6 (1916) | 4720 |
| Number of conditions median (IQR) | 1 (2) | 1 (2) | 4720 |
| Current drinker | 63.4 (2807) | 63.3 (2894) | 4570 |
| Self-rated over- or underweight | 58.0 (2567) | 57.5 (2695) | 4691 |
| Self-isolation* | 32.5 (1441) | 32.7 (1544) | 4720 |
| Close contact with people who tested positive* | 13.8 (609) | 13.3 (629) | 4716 |

* Data was collected in ELSA COVID-19 Substudy

**Figure S1. Dendrogram of cluster analysis**

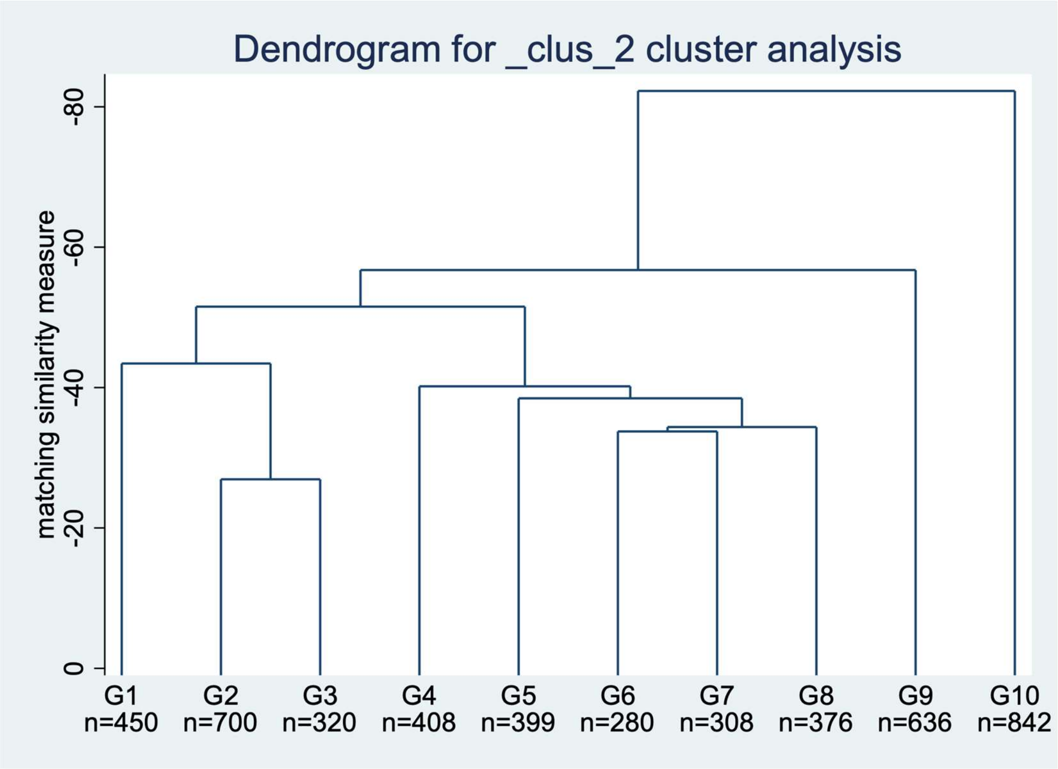
